# Pancreas Procurement from Deceased Donors for Islet Cells: Updated Data and Gaps in Reporting

**DOI:** 10.1101/2025.02.21.25322017

**Authors:** David S Goldberg, Erin Tewksbury, Matthew Wadsworth

**Affiliations:** Division of Digestive Health and Liver Diseases, University of Miami Miller School of Medicine, Miami, FL; Life Connection of Ohio, Toledo, OH

**Author notes:** Corresponding author: David Goldberg, MD, MSCE, Don Soffer Clinical Research Building 1120 NW 14^th^ Street, Room 807, Miami, FL 33136. **AUTHORSHIP PAGE**. **DG:** Participated in research design, data analysis, writing of the paper, final manuscript approval. **ET:** Participated in research design, data analysis, writing of the paper, final manuscript approval. **MW:** Participated in research design, data analysis, writing of the paper, final manuscript approval. **Disclosure:** The authors declare no conflicts of interest. **Funding:** None.

## Abstract

**Background:** In 2020, CMS updated the ‘Final Rule’ for organ procurement organizations (OPO), the non-profit organizations tasked with managing organ donation. The final rule included new metrics of performance with a carveout whereby pancreata procured for research count as a “donor” to reward OPOs procuring pancreata for islet cell research or transplantation. Yet data suggested OPOs took advantage of this loophole, leading to investigations by the Senate Finance Committee and updated guidance in 2024, to allow OPOs to revise the coding of these data.

**Methods:** We evaluated OPTN data on OPO reporting of research pancreata procurements and compared the originally reported data to data reported with the new codes. We then compared these data to data from the Integrated Islet Cell Distribution Program (IIDP) and the Network for Pancreatic Organ Donors with Diabetes (nPOD) to quantify the magnitude of the differences in OPO reported data and data on donor pancreata utilization from the two largest NIH networks.

**Results:** Although more than half of the OPOs adjusted their data using the new codes, 8 continued to report large numbers of pancreata procured for research. Despite this, there was a nearly 20-fold difference between what OPOs reported as procured for research compared to what was used by the IIDP and nPOD.

**Conclusions:** Concerns remain about the veracity of reporting of procurement of research pancreata from deceased donors, necessitating oversight and audits from CMS and other regulatory agencies.

## INTRODUCTION

In November 2020, the Centers for Medicare and Medicaid Services (CMS) updated the ‘Final Rule’ for organ procurement organizations (OPO) in 2020 to include new metrics of organ procurement organization (OPO) performance.^1^ In addition to implementing a new metric using administrative data, CMS included a carveout whereby pancreata procured for research would count as a “donor” (or donor organ). The rationale for this carveout was to give OPOs credit for procuring pancreata for to be used for islet cell research or islet cell transplantation.^1^ Following the implementation of this rule, there was a massive increase in the number of reported procurements of research pancreata.^2–4^ As a result, the Senate Finance Committee began an ongoing investigation in March 2023 due to concerns that these pancreata were not being procured for “legitimate research purposes,” in contradiction to the final rule which specified that, “only bona fide research conducted by a qualified researcher using a pancreas from an organ donor” would be counted in the metric.^1,5,6^ This investigation is ongoing, with the Senate Finance Committee still awaiting responses from 10 OPOs who received letters related to their research pancreata procurements, the protocols they were used for, along with other information.^5,6^

CMS updated its OPO guidance in 8/2024 and added language stating, “Pancreata will be considered ‘used’ for research if they are *accepted for use* in bona fide research conducted by a qualified researcher, such as those approved by the National Institutes of Health.^7^ Furthermore, in conjunction with the Health Resources and Services Administration (HRSA) and the Organ Procurement and Transplantation Network (OPTN), new codes were developed to allow OPOs to, “differentiate between pancreata used for islet cell transplantation or research and pancreata used for non-islet cell research.”^7^ These new codes specified whether a pancreas was recovered for research and either, “Sent for non-islet cell research” or “Accepted for islet cell research.”^7^ These new codes took effect for 2024, and OPOs were mandated to recode their data on recovered pancreata from 2021-2023.

The updated data on research pancreata was recently made available by the OPTN. Therefore, we sought to evaluate changes in research pancreata procurement based on updated coding and to determine whether there were OPO-level differences in their reported data. Furthermore, we sought to evaluate these updated OPTN data to publicly available data on pancreas and islet cell research in order to quantify the magnitude in the difference between reported research pancreata procurements and islet cell research from the two largest National Institutes of Health (NIH) funded networks focused on islet cell research from organ donors.

## METHODS

### Research pancreata procurements

We conducted a retrospective cohort study following the STROBE reporting guidelines. We evaluated (OPTN) data of reported research pancreata from 1/1/2018-12/31/2024. Although the data from 1/1/2018-12/31/2020 did not undergo recording using the new research pancreata procurement codes, these earlier data were used to contextualize what was previously reported to the OPTN compared to data from the two largest NIH-funded islet cell research networks. We compared national-and OPO-level data on research pancreas procurements using the original and new codes on research pancreata procurements.

### Islet cell research

The process of isolating islet cells for from deceased donors is a complex and costly process that requires processing and isolation of fresh donor pancreata. This process is costly and requires a highly skilled islet cell isolation team with specialized equipment. The limited supply and the limited expertise in islet cell isolation was a longstanding barrier to islet cell isolation in the US.^8–12^ In 2004, the National Institutes of Diabetes and Digestive and Kidney Diseases (NIDDK) begin to fund the Integrated Islet Cell Distribution Program (IIDP) to oversee the distribution of human islets for diabetes research.^9,13^ This network serves to isolate and distribute islet cells for a broad array of research that requires the complex and costly process of islet cells to be isolated as a first step. Due to the costs of expertise of islet cell isolation, in the US, the process of islet cell isolation has almost exclusively been limited to these centers. As an example, in 2009, there were only 8 academic institutions in the entire United States that were active and funded to provide process and provide islet cells for research.^9^ The IIDP originally included five centers, and over the last few years has expanded to 10 centers across the country.^13^ The IIDP has publicly available data on its sources of research pancreases from 2018-2024, categorized nationally and per OPO. These data were used to capture data on research pancreases used by the largest group of islet cell isolation centers in the US.

The other major source of islet cell research in the US is the NIH-funded islet cell research through the Network for Pancreatic Organ Donors with Diabetes (nPOD).^14^ The nPOD is focused on research related to type 1 diabetes, and studies pancreata (including islet cells) from deceased donors with type 1 diabetes, non-diabetic donors with positive diabetes autoantibodies, and non-diabetic autoantibody negative controls. Although the nPOD does not involve islet cell isolation like the IIDP, the deceased donors analyzed and distributed by the nPOD involve analyses of islet cells, and therefore meets CMS criteria of “bona fide research conducted by a qualified researcher.”^7,13^ We evaluated publicly available data from the nPOD, which were only available as an aggregate number of donor pancreata from 2017-2024.

We did not include data from the International Institute for the Advancement of Medicine (IIAM) because they do not provide publicly available data on pancreata from which islet cells were accepted for research, and the only research projects related to pancreata or islet cells listed by the IIAM are those involving the nPOD.^15^

This study was considered exempt from Institutional Review Board review as it involved non-human subjects research of publicly available deidentified data.

## RESULTS

### Changes in reporting of research pancreata procurements

As previously reported, based on the original coding of research pancreata procurements, there was a rapid rise beginning in 2021, following the publication of the CMS OPO final rule, which increased six-fold in 2023 relative to 2020 (Figure 1).^2–4^ However, based on the research pancreata codes, the increase in the number of reported research pancreata procurement began in 2022, rather than 2021 and was nearly half previously reported. For example, in 2023, the number of procured research pancreata decreased from an initially reported value of 3,338 to 1,812 when restricted to those OPOs reported were accepted for islet cell research.

**Figure 1:**
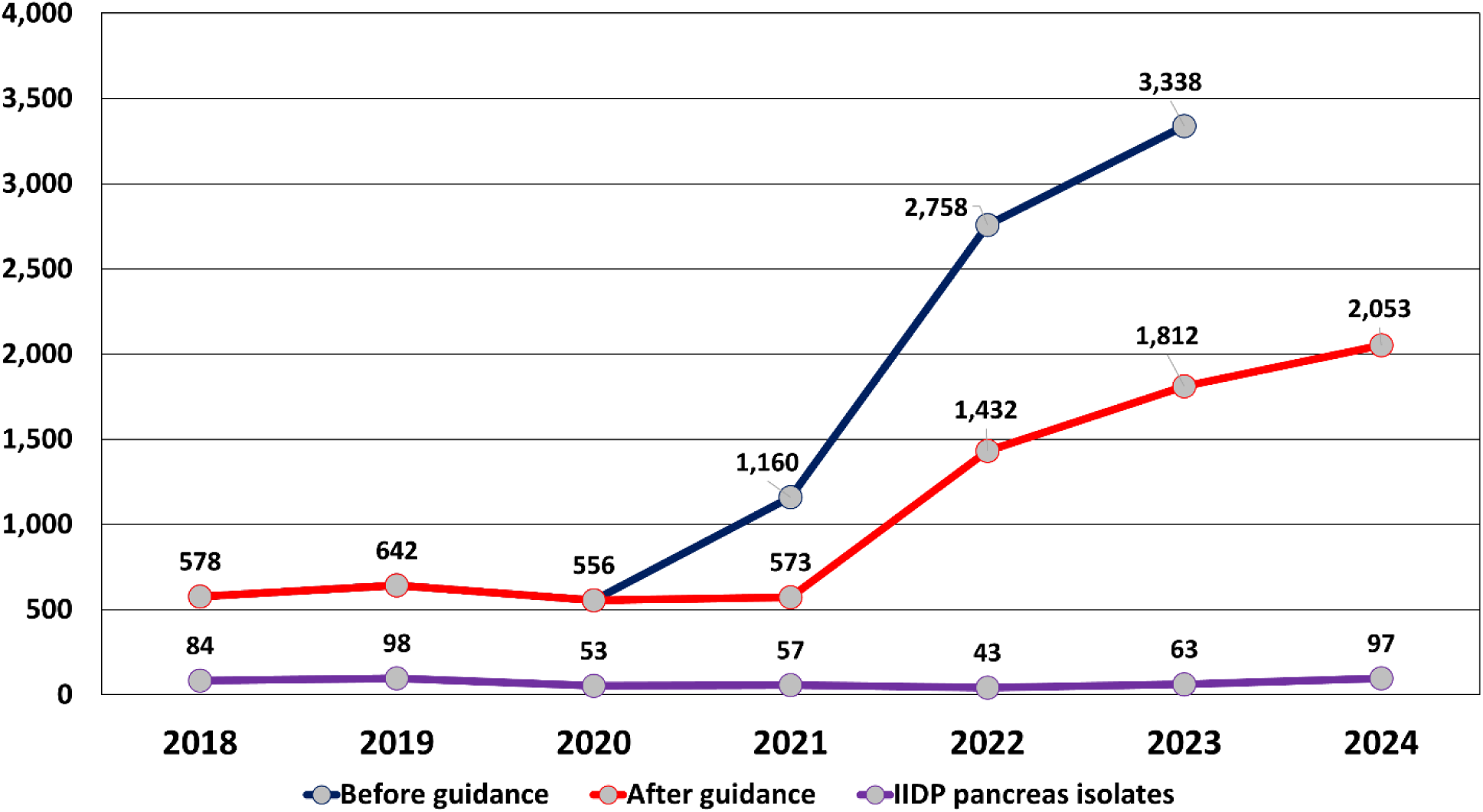
Annual data on research pancreas procurements compared to IIDP-reported pancreata involved in islet cell isolation, 2018-2024

The changes in the reporting of the number of research pancreata using the new codes was variable across OPOs (Figures 2a-2c, Table 1), without a specific geographic pattern or correlation with the original volume of reported research pancreata procurements. For example, in 2023, there were 12 OPOs that originally reported procuring more than 100 research pancreata. However, after CMS requested OPOs revise their data reporting using the new codes, five of these OPOs reported more than 100 fewer research pancreata procurement, most notably LifeQuest Organ Recovery Services (FLUF) that decreased their reported research pancreata procurements from 230 to 2, and Legacy of Hope (ALOB) that decreased their reported research pancreata procurements from 226 to 0. Conversely, six of these OPOs reported a decrease of fewer than 30 research pancreata procurements based on the updated coding, most notably OneLegacy (CAOP) that only decreased their number of research pancreata procurements from 520 to 492 (Table 1).

**Figure 2.**
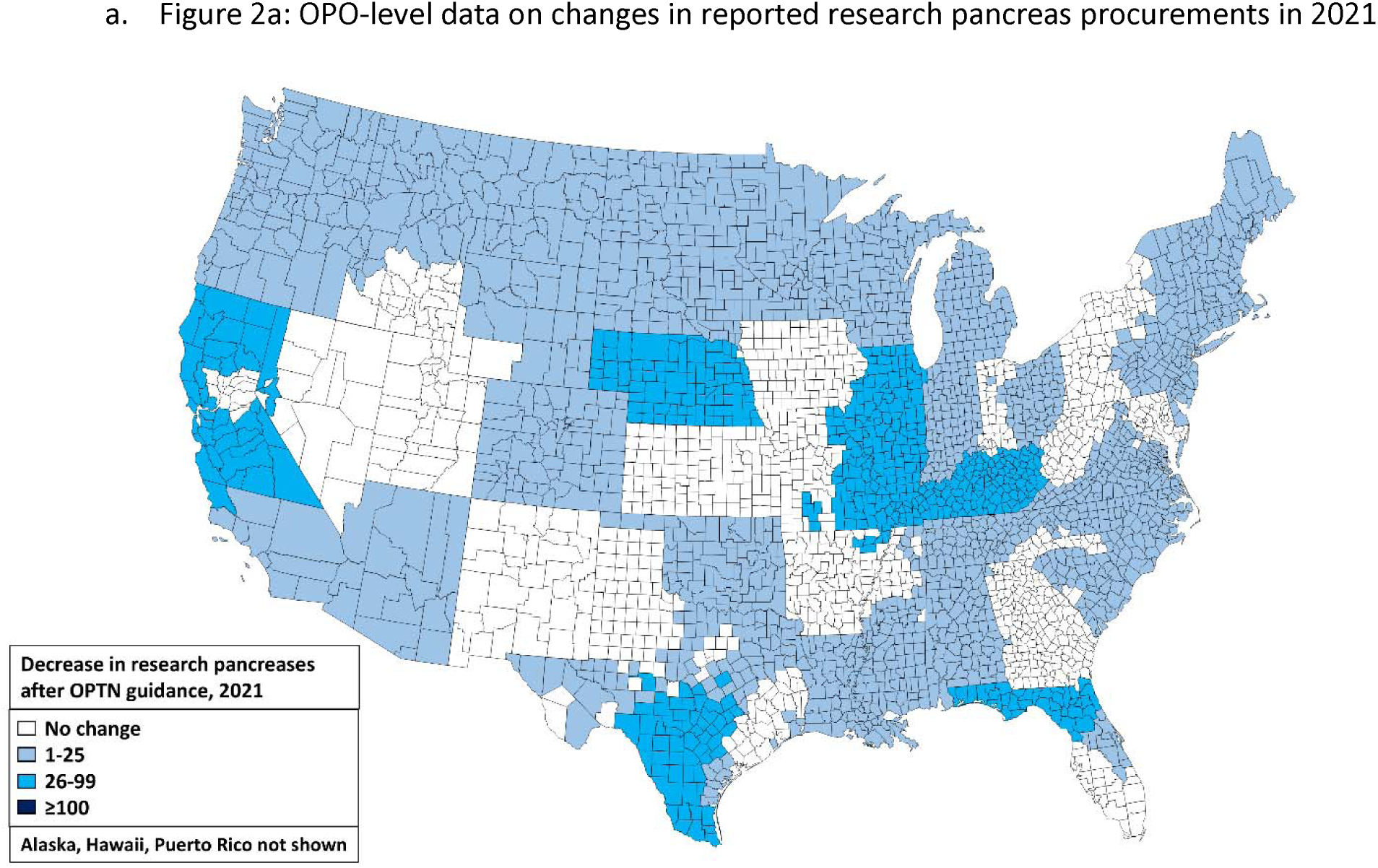
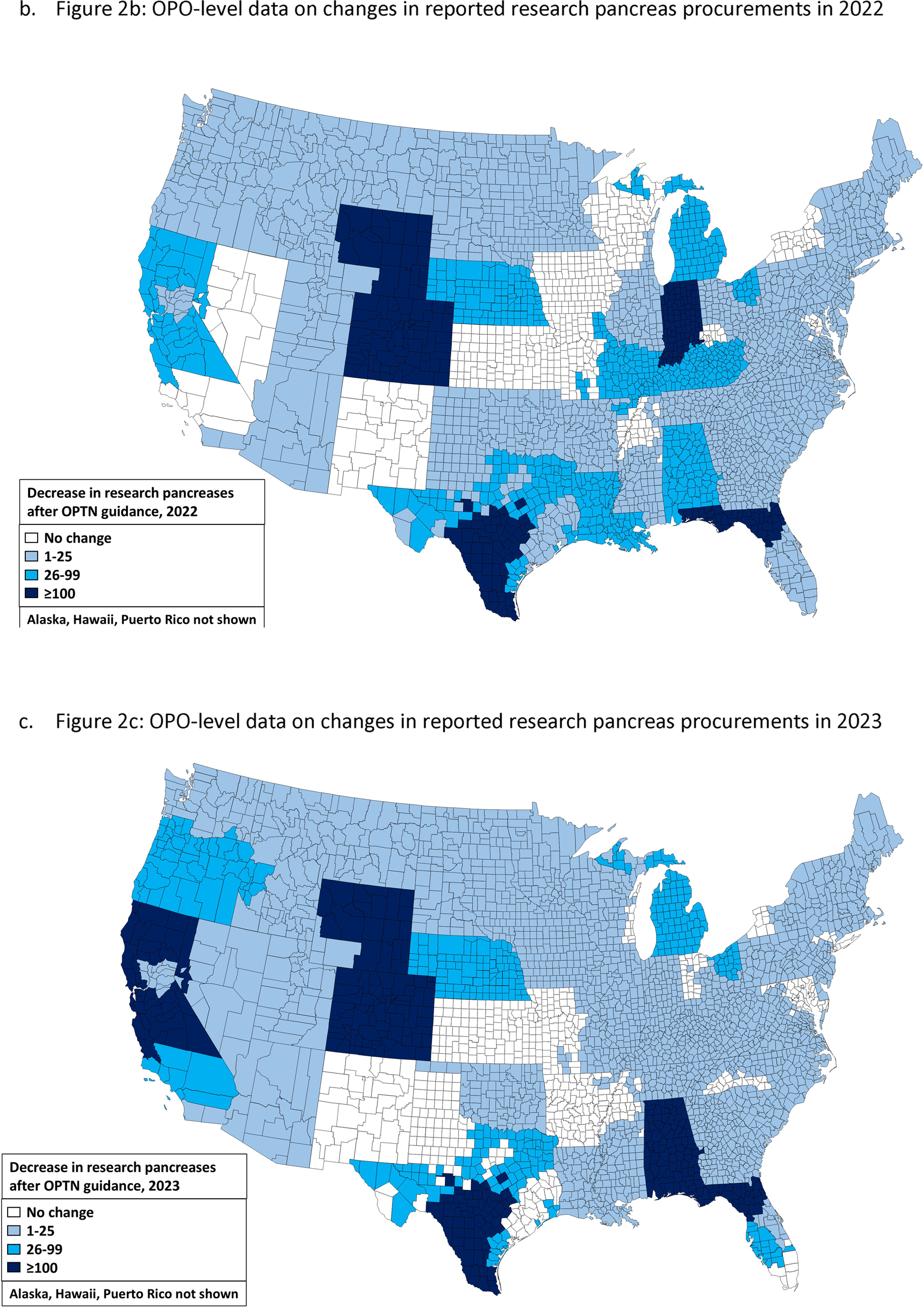
(three panels): OPO-level data on changes in reported research pancreata procurements

**Table 1:** Data on reported research pancreata before and after CMS 8/2024 guidance.

|  | Research pancreata<br>before guidance |  |  | Research pancreata after<br>guidance |  |  | Δ research pancreata after<br>guidance |  |  |
| --- | --- | --- | --- | --- | --- | --- | --- | --- | --- |
| OPO | 2021 | 2022 | 2023 | 2021 | 2022 | 2023 | 2021 | 2022 | 2023 |
| ALOB | 9 | 59 | 226 | 1 | 0 | 0 | -8 | -59 | -226 |
| AROR | 0 | 1 | 0 | 0 | 0 | 0 | 0 | -1 | 0 |
| AZOB | 26 | 83 | 161 | 18 | 76 | 155 | -8 | -7 | -6 |
| CADN | 68 | 194 | 182 | 8 | 118 | 60 | -60 | -76 | -122 |
| CAGS | 4 | 4 | 2 | 4 | 3 | 1 | 0 | -1 | -1 |
| CAOP | 90 | 441 | 520 | 83 | 441 | 492 | -7 | 0 | -28 |
| CASD | 3 | 21 | 11 | 1 | 0 | 7 | -2 | -21 | -4 |
| CORS | 2 | 130 | 148 | 1 | 1 | 11 | -1 | -129 | -137 |
| FLFH | 28 | 68 | 56 | 21 | 57 | 37 | -7 | -11 | -19 |
| FLMP | 2 | 6 | 6 | 2 | 5 | 6 | 0 | -1 | 0 |
| FLUF | 98 | 176 | 230 | 4 | 0 | 2 | -94 | -176 | -228 |
| FLWC | 1 | 11 | 48 | 1 | 3 | 0 | 0 | -8 | -48 |
| GALL | 2 | 21 | 29 | 2 | 12 | 6 | 0 | -9 | -23 |
| HIOP | 0 | 0 | 0 | 0 | 0 | 0 | 0 | 0 | 0 |
| IAOP | 0 | 0 | 7 | 0 | 0 | 0 | 0 | 0 | -7 |
| ILIP | 63 | 44 | 30 | 11 | 29 | 13 | -52 | -15 | -17 |
| INOP | 17 | 144 | 12 | 2 | 12 | 2 | -15 | -132 | -10 |
| KYDA | 105 | 114 | 151 | 64 | 67 | 126 | -41 | -47 | -25 |
| LAOP | 4 | 35 | 11 | 3 | 2 | 3 | -1 | -33 | -8 |
| MAOB | 4 | 20 | 21 | 3 | 3 | 7 | -1 | -17 | -14 |
| MDPC | 0 | 1 | 5 | 0 | 0 | 5 | 0 | -1 | 0 |
| MIOP | 13 | 36 | 76 | 2 | 3 | 3 | -11 | -33 | -73 |
| MNOP | 8 | 12 | 19 | 0 | 2 | 3 | -8 | -10 | -16 |
| MOMA | 138 | 215 | 240 | 61 | 173 | 220 | -77 | -42 | -20 |
| MSOP | 22 | 15 | 4 | 0 | 2 | 0 | -22 | -13 | -4 |
| MWOB | 3 | 1 | 2 | 3 | 1 | 2 | 0 | 0 | 0 |
| NCCM | 50 | 67 | 45 | 50 | 66 | 45 | 0 | -1 | 0 |
| NCNC | 4 | 7 | 30 | 2 | 6 | 24 | -2 | -1 | -6 |
| NEOR | 37 | 55 | 26 | 0 | 0 | 0 | -37 | -55 | -26 |
| NJTO | 44 | 84 | 116 | 26 | 73 | 101 | -18 | -11 | -15 |
| NMOP | 0 | 0 | 0 | 0 | 0 | 0 | 0 | 0 | 0 |
| NVLV | 4 | 1 | 21 | 4 | 1 | 1 | 0 | 0 | -20 |
| NYAP | 1 | 2 | 9 | 0 | 0 | 2 | -1 | -2 | -7 |
| NYFL | 0 | 0 | 6 | 0 | 0 | 2 | 0 | 0 | -4 |
| NYRT | 37 | 41 | 54 | 36 | 40 | 54 | -1 | -1 | 0 |
| NYWN | 0 | 0 | 0 | 0 | 0 | 0 | 0 | 0 | 0 |
| OHLB | 2 | 63 | 172 | 1 | 29 | 127 | -1 | -34 | -45 |
| OHLC | 0 | 3 | 2 | 0 | 2 | 2 | 0 | -1 | 0 |
| OHLP | 13 | 64 | 114 | 1 | 40 | 110 | -12 | -24 | -4 |
| OHOV | 41 | 48 | 72 | 41 | 48 | 70 | 0 | 0 | -2 |
| OKOP | 5 | 24 | 4 | 1 | 1 | 3 | -4 | -23 | -1 |
| ORUO | 26 | 30 | 47 | 10 | 8 | 3 | -16 | -22 | -44 |
| PADV | 23 | 19 | 21 | 20 | 17 | 19 | -3 | -2 | -2 |
| PATF | 29 | 33 | 34 | 29 | 30 | 33 | 0 | -3 | -1 |
| PRLL | 2 | 35 | 83 | 0 | 0 | 0 | -2 | -35 | -83 |
| SCOP | 4 | 1 | 3 | 3 | 0 | 0 | -1 | -1 | -3 |
| TNDS | 2 | 4 | 4 | 0 | 3 | 3 | -2 | -1 | -1 |
| TNMS | 1 | 3 | 0 | 1 | 3 | 0 | 0 | 0 | 0 |
| TXGC | 7 | 10 | 4 | 7 | 6 | 4 | 0 | -4 | 0 |
| TXSA | 55 | 174 | 192 | 2 | 1 | 3 | -53 | -173 | -189 |
| TXSB | 8 | 66 | 38 | 5 | 10 | 10 | -3 | -56 | -28 |
| UTOP | 4 | 7 | 7 | 4 | 1 | 3 | 0 | -6 | -4 |
| VATB | 1 | 3 | 2 | 0 | 0 | 0 | -1 | -3 | -2 |
| WALC | 6 | 5 | 6 | 1 | 0 | 4 | -5 | -5 | -2 |
| WIDN | 9 | 22 | 7 | 2 | 9 | 7 | -7 | -13 | 0 |
| WIUW | 27 | 19 | 22 | 25 | 19 | 21 | -2 | 0 | -1 |

**Table 2:** Number of reported pancreata procured from deceased donors for islet cell research and in parentheses the number of pancreata with islet cells isolated by IIDP*.

|  | 2018 | 2019 | 2020 | 2021 | 2022 | 2023 | 2024 |
| --- | --- | --- | --- | --- | --- | --- | --- |
| ALOB | 1 (0) | 4 (0) | 0 (0) | 1 (0) | 0 (0) | 0 (0) | 0 (0) |
| AROR | 0 (0) | 1 (0) | 1 (0) | 0 (0) | 0 (0) | 0 (0) | 0 (0) |
| AZOB | 12 (0) | 25 (1) | 21 (4) | 18 (6) | 76 (3) | 155 (0) | 193 (3) |
| CADN | 18 (3) | 27 (1) | 15 (2) | 8 (2) | 118 (0) | 60 (0) | 69 (1) |
| CAGS | 4 (0) | 3 (0) | 3 (0) | 4 (0) | 3 (0) | 1 (0) | 2 (0) |
| CAOP | 106 (34) | 82 (41) | 43 (21) | 83 (28) | 441 (20) | 492 (26) | 453 (32) |
| CASD | 1 (0) | 17 (2) | 8 (0) | 1 (0) | 0 (3) | 7 (2) | 1 (2) |
| CORS | 2 (0) | 1 (0) | 2 (0) | 1 (0) | 1 (0) | 11 (1) | 2 (0) |
| CTOP | 1 (0) | 0 (0) | 1 (0) | NA | NA | NA | NA |
| DCTC | 13 (0) | 14 (0) | 14 (0) | 7 (0) | 9 (0) | NA | NA |
| FLFH | 10 (0) | 10 (0) | 0 (0) | 21 (0) | 57 (0) | 37 (0) | 22 (0) |
| FLMP | 2 (1) | 2 (1) | 0 (0) | 2 (2) | 5 (0) | 6 (1) | 10 (5) |
| FLUF | 4 (0) | 1 (0) | 5 (0) | 4 (0) | 0 (0) | 2 (0) | 1 (0) |
| FLWC | 17 (0) | 41 (0) | 18 (0) | 1 (0) | 3 (0) | 0 (0) | 3 (0) |
| GALL | 6 (0) | 6 (0) | 3 (0) | 2 (0) | 12 (0) | 6 (0) | 5 (10) |
| HIOP | 0 (0) | 0 (0) | 0 (0) | 0 (0) | 0 (0) | 0 (0) | 0 (0) |
| IAOP | 0 (0) | 0 (0) | 1 (0) | 0 (0) | 0 (0) | 0 (0) | 0 (0) |
| ILIP | 3 (0) | 11 (0) | 28 (0) | 11 (1) | 29 (2) | 13 (0) | 2 (1) |
| INOP | 2 (0) | 9 (0) | 5 (0) | 2 (2) | 12 (0) | 2 (0) | 137 (3) |
| KYDA | 31 (0) | 27 (0) | 71 (0) | 64 (0) | 67 (0) | 126 (1) | 178 (2) |
| LAOP | 4 (0) | 6 (0) | 3 (0) | 3 (0) | 2 (0) | 3 (1) | 178 (0) |
| MAOB | 9 (0) | 2 (0) | 3 (0) | 3 (0) | 3 (0) | 7 (0) | 8 (1) |
| MDPC | 1 (0) | 3 (0) | 2 (0) | 0 (0) | 0 (0) | 5 (0) | 10 (1) |
| MIOP | 19 (0) | 6 (0) | 1 (0) | 2 (0) | 3 (0) | 3 (0) | 4 (0) |
| MNOP | 5 (0) | 2 (0) | 1 (0) | 0 (0) | 2 (0) | 3 (0) | 4 (0) |
| MOMA | 5 (0) | 20 (0) | 85 (0) | 61 (0) | 173 (0) | 220 (0) | 234 (0) |
| MSOP | 1 (0) | 0 (0) | 12 (0) | 0 (0) | 2 (0) | 0 (0) | 3 (0) |
| MWOB | 7 (0) | 6 (0) | 3 (0) | 3 (0) | 1 (0) | 2 (0) | 45 (0) |
| NCCM | 2 (0) | 0 (0) | 1 (0) | 50 (0) | 66 (0) | 45 (0) | 40 (0) |
| NCNC | 22 (1) | 13 (0) | 6 (0) | 2 (0) | 6 (0) | 24 (1) | 26 (1) |
| NEOR | 23 (0) | 29 (0) | 14 (0) | 0 (0) | 0 (0) | 0 (0) | 0 (0) |
| NJTO | 1 (0) | 1 (0) | 23 (0) | 26 (0) | 73 (0) | 101 (0) | 46 (0) |
| NMOP | 0 (0) | 1 (0) | 0 (0) | 0 (0) | 0 (0) | 0 (0) | 0 (0) |
| NVLV | 3 (0) | 5 (0) | 7 (1) | 4 (1) | 1 (0) | 1 (0) | 3 (1) |
| NYAP | 12 (0) | 3 (0) | 3 (0) | 0 (0) | 0 (0) | 2 (0) | 1 (0) |
| NYFL | 1 (0) | 0 (0) | 0 (0) | 0 (0) | 0 (0) | 2 (0) | 1 (0) |
| NYRT | 45 (0) | 30 (0) | 23 (0) | 36 (0) | 40 (0) | 54 (0) | 35 (0) |
| NYWN | 1 (0) | 0 (0) | 0 (0) | 0 (0) | 0 (0) | 0 (0) | 0 (0) |
| OHLB | 3 (0) | 1 (0) | 1 (0) | 1 (0) | 29 (0) | 127 (0) | 91 (0) |
| OHLC | 2 (0) | 2 (0) | 3 (0) | 0 (0) | 2 (0) | 2 (0) | 4 (0) |
| OHLP | 0 (0) | 4 (0) | 1 (0) | 1 (0) | 40 (0) | 110 (1) | 87 (5) |
| OHOV | 2 (0) | 0 (0) | 1 (0) | 41 (0) | 48 (0) | 70 (0) | NA (0) |
| OKOP | 6 (0) | 1 (0) | 1 (0) | 1 (0) | 1 (0) | 3 (0) | 20 (0) |
| ORUO | 15 (0) | 62 (0) | 13 (0) | 10 (1) | 8 (1) | 3 (0) | 3 (0) |
| PADV | 18 (1) | 18 (10) | 24 (4) | 20 (9) | 17 (4) | 19 (2) | 28 (12) |
| PATF | 21 (0) | 25 (0) | 15 (0) | 29 (0) | 30 (4) | 33 (11) | 50 (7) |
| PRLL | 1 (0) | 3 (0) | 2 (0) | 0 (0) | 0 (0) | 0 (0) | 0 (0) |
| SCOP | 5 (0) | 8 (0) | 0 (0) | 3 (0) | 0 (0) | 0 (0) | 0 (0) |
| TNDS | 3 (0) | 5 (0) | 0 (0) | 0 (0) | 3 (0) | 3 (0) | 3 (0) |
| TNMS | 1 (0) | 1 (0) | 2 (0) | 1 (0) | 3 (0) | 0 (0) | 2 (0) |
| TXGC | 17 (1) | 16 (2) | 10 (0) | 7 (0) | 6 (0) | 4 (0) | 2 (2) |
| TXSA | 13 (1) | 7 (0) | 4 (1) | 2 (0) | 1 (0) | 3 (0) | 2 (0) |
| TXSB | 9 (0) | 12 (0) | 5 (0) | 5 (0) | 10 (0) | 10 (1) | 3 (1) |
| UTOP | 3 (0) | 3 (0) | 3 (0) | 4 (0) | 1 (0) | 3 (0) | 7 (0) |
| VATB | 24 (0) | 16 (0) | 6 (0) | 0 (0) | 0 (0) | 0 (0) | 1 (0) |
| WALC | 5 (0) | 25 (0) | 26 (0) | 1 (0) | 0 (0) | 4 (4) | 0 (0) |
| WIDN | 3 (1) | 2 (1) | 0 (0) | 2 (0) | 9 (0) | 7 (3) | 7 (2) |
| WIUW | 33 (2) | 23 (9) | 13 (6) | 25 (1) | 19 (2) | 21 (6) | 27 (9) |
| National | 578 (84) | 642 (98) | 556 (53) | 573 (57) | 1,432 (43) | 1,812 (63) | 2,053 (97) |

\* Number of research pancreata reported by OPOs to the OPTN. Data from 2021-2024 are based on updated coding where OPO report pancreata “accepted for islet cell research.” Data on pancreata with islet cells isolated by the IIDP shown in parentheses and publicly available from the IIDP.<sup>8,9</sup>

### Comparing OPTN to IIDP and nPOD data

From 2018-2024, the number of donor research pancreata from which islet cells were isolated from the IIDP ranged from 43 to 97 (495 total over a 7-year period; Figure 1). The nPOD, which only reports aggregate data, reported a total of 688 deceased donors procured for research within the nPOD from 2007-2024. Together, based on seven years of IIDP data and 18 years of nPOD data, a total of 1,183 research pancreata were used by qualified researchers across these two large multi-center NIDDK funded networks, less than the number OPOs reported were accepted for islet cell research in 2022 alone. In 2024, procured research pancreata were concentrated in 8 OPOs, who reported collecting 1,551 pancreata “accepted for islet” cell research, 75.6% of national data. However, the IIDP, with its 10 islet cell isolation centers, only collected 45 pancreata for islet cell isolates from these OPOs (Table 1).^13^

The gap between reported research pancreata procurements and IIDP data increased substantially after publication of the new CMS final rule in 11/2020. From 2018-2021, the first year during which the new OPTN codes were used for research pancreata procurement, OPOs reported procuring pancreata for islet cell research that was 6-10 times higher than those isolated by the IIDP, with a 10-fold difference in 2021.^13^ Yet from 2022-2024, this discrepancy grew to a 20-30 fold difference, despite the IIDP adding five centers during this period (Figure 1).^9,13^ And this gap between OPO-reported data and IIDP data was not uniform across OPOs, and grew over time. For example, LifeBanc (OHLB) reported a total of 6 research pancreata procurements to the OPTN between 2018-2021, and zero pancreata were isolated by the IIDP among donors from LifeBanc. Yet from 2022-2024, LifeBanc reported to the OPTN that it procured 29, 127, and 91 research pancreata that were accepted for islet cell research; yet the IIDP did not isolate islet cells from a single pancreas donor from this OPO (Table 1). Additionally, the IIDP isolated islet cells from approximately 20-40 pancreata per year from 2018-2024 from OneLegacy (CAOP), yet despite these stable numbers, OneLegacy reported to the OPTN a more than 5-fold increase in the number of research pancreata accepted for islet cell research (from 83 to more than 400; Table 1).

Similar large gaps were seen among other OPOs including Donor Network of Arizona (AZOB), Donor Network West (CADN), Kentucky Organ Donor Affiliates (KYDA), Indiana Donor Network (INOP), Louisiana Organ Procurement Agency (LAOP), Mid-America Transplant Services (MOMA), and Lifeline of Ohio (OHLP).

## DISCUSSION

In response to the CMS OPO final rule of 2020, OPOs began to report a marked increase in the number of research pancreata procurements. In response to Senate Finance Committee Investigations, and peer-reviewed data raising concerns about these findings, CMS updated their guidance, resulting in a continued, albeit attenuated growth in research pancreata. However, the changes in reporting of data was not uniform across OPOs, with some revising their data to reflect only rare procurement of research pancreata, while others continuing to report high rates of research pancreata procurement. Despite increased stringency from CMS about the definition of a research pancreas procurement, the gap between the number of research pancreata OPOs report have been accepted from islet cell research is an order of magnitude higher than what the two largest NIH consortium focused on donor islet cell research have reported. Despite inquiries from the Senate Finance Committee, the disposition of the remainder of these islet cells remains unclear, with few other qualified centers in the US available to process and isolate these ilset cells for research.

The updated CMS guidance specify that research pancreata must be associated with islet cells accepted for islet cell research or islet cell transplant. Islet cell transplantation is overseen by the Food and Drug Administration, and remains rare, with fewer than 20 cases per year, far too few to explain the gap between IIDP and nPOD data and that reported by OPOs. IIDP represented the leading iselt cell islet cell isolation centers in the US, and despite expanding from 5 to 10 isolation centers, the divide between what has been reported as research pancreata accepted for islet cell research and pancreata with islet cells isolated by the IIDP has only grown. And although the nPOD does not report annualized data, with 688 pancreata used for research (average 38 per year) over 18 years, even with increases over time, the donor research pancreata supplied by this NIDDK-funded network would minimally bridge the gap between OPO-reported versus IIDP/nPOD-reported data. A critical point is that while the nPOD is a specific research network focused on diabetes, the IIDP is a distribution program, whereby the NIDDK funds the leading isolation centers to isolate and distribute iselt cells for research. While there is the potential that over the last several years OPOs have developed new scientific collaborations to stimulate islet cell research, given the cost, expertise, and requirement for specialized equipment and specialized isolation teams, it is implausible that a sufficient amount islet cell isolation could have occurred outside of the IIDP to explain the gap in what OPOs report versus what is captured by the IIDP and nPOD.^8–12^

We acknowlege there are islet cell isolation centers that may not be associated with the IIDP, and other networks that may assist in procuring research pancreata for islet cell research. Yet the IIDP and nPOD reflect the two largest federally funded consortium focused on this. And given the cost, logistics, and expertise needed for islet cell isolation, it remains implausible to expect there are enough centers not captured by the IIDP or nPOD supplying islet cells for research to “qualified researchers.” The Senate Finance Committee has been investigating this issue for nearly two years, and have yet to report on data from OPOs that help to elucidate the disposition of these research pancreata they report are accepted for islet cell research.

Procurement of research pancreata uses taxpayer dollars, could potentially be linked to CMS cost reports, and most importantly, involves procuring organs from deceased donors. Increasing research in diabetes and other pancreatic conditions is critical, but so to is the appropriate procurement of organs, and accurate categorization of them as being used for “islet cell research.” It is critical that CMS and other regulatory agencies audit OPOs about the disposition of research pancreata to ensure data are reported accurately as this has implications for CMS metrics, CMS expenditures of organ donation, and the public trust in organ donation. Organ donation and transplantation is highly regulated, and the disposition of organs that are transplanted is recorded. A similar process could be followed whereby OPOs report the disposition (i.e., research entity/organization/researcher) of research pancreata.

## Abbreviations

CMS: Centers for Medicare and Medicaid Services
OPOs: Organ procurement organizations
HRSA: Health Resources and Services Administration
OPTN: Organ Procurement and Transplantation Network
IIDP: Integrated Islet Cell Distribution Program
nPOD: Network for Pancreatic Organ Donors with Diabetes

## Data Availability

The OPTN data are available by request from the OPTN, which were made available in aggregate data per OPO per year, and due to the DUA require other investigators request the data from the OPTN. the IIDP data are available on the IIDP website: https://iidp.coh.org/Overview/Program-Statistics

## Acknowledgments

This work was supported in part by Health Resources and Services Administration contract HHSH250-2019-00001C. The content is the responsibility of the authors alone and does not necessarily reflect the views or policies of the Department of Health and Human Services, nor does mention of trade names, commercial products, or organizations imply endorsement by the U.S. Government. Data on islet cell research from the IIDP were obtained from publicly available data online from the IIDP. The opinions and perspectives of these data are our own. Dr. Goldberg and Erin Tewksbury had full access to all the data in the study and takes responsibility for the integrity of the data and the accuracy of the data analysis. There was no funding organization or sponsor involved in the design and conduct of the study; collection, management, analysis, and interpretation of the data; preparation, review, or approval of the manuscript; and decision to submit the manuscript for publication.

